# Risk Factors for Admission into COVID-19 General Wards, Sub-Intensive and Intensive Care Units among SARS-CoV-2 Positive Subjects in the Municipality of Bologna, Italy

**DOI:** 10.1101/2023.07.12.23292559

**Authors:** Sofia Raponi, Francesco Durazzi, Nicolas Riccardo Derus, Enrico Giampieri, Rossella Miglio, Gastone Castellani, Claudia Sala, with the Bologna MODELS4COVID Study Group

## Abstract

This is a retrospective cohort study aimed at identifying the risk factors for the hospitalization of patients with COVID-19 in the municipality of Bologna. A total of 32500 patients that tested positive for COVID-19 from February 28/2020 to October 13/2021 in the municipality of Bologna were included. The Kaplan-Meier method was used to estimate changes during time of ICU hospitalization for all patients as well as stratifying subjects by sex. A multi-state Cox’s proportional hazard model was fitted to investigate predictors of ICU and non-ICU hospitalization. Age, sex, calendar period of diagnosis, comorbidities and vaccination status of patients at the time of diagnosis were considered as candidate predictors. In general, male sex and advanced age resulted to be poor prognostic factors of COVID-19 outcomes. An exception was found for the over 80 age group which showed a decrease in the risk of ICU hospitalization compared to 70-79 (HR 0.57 95% CI 0.36 - 0.90 for DIAG*→*ICU; HR 0.40 95% CI 0.28 - 0.58 for HOSP*→*ICU). Having contracted the disease during the first wave was associated with a significant greater risk of hospitalization than during the second wave, while no difference in the risk of ICU admission was found between the second and third waves. Fully immunized patients showed a significant decrease in the risk of ICU and non-ICU hospitalization compared to the unvaccinated patients (HR 0.23 95% CI 0.16 - 0.31 for DIAG*→*HOSP; HR 0.10 95% CI 0.01 - 0.73 for DIAG*→*ICU). Chronic kidney failure and asthma were risk factors for non-ICU hospitalization. Diabetes and embolism were risk factors for both direct ICU and non-ICU hospitalization. The study of factors associated with a negative course of the COVID-19 disease allows to prevent fatal outcomes, establish priorities in the treatment of the disease and improve the management of hospital resources and the pandemic itself.

## Introduction

Since the beginning of 2020, the COVID-19 pandemic has become a threat to global health. The SARS-CoV-2 virus that causes the COVID-19 disease was first found in China in late 2019 [1]. Since that time it has spread fast all over the globe and Italy has become one of the European most affected countries [2].

The rapid diffusion of this new virus and the unpredictability of the disease course has led to the development of numerous studies aimed at investigating prognostic factors of COVID-19 outcomes in order to predict the severity of the disease. These descriptive studies allow researchers and doctors to early identify patients most at risk of disease aggravation and policy makers to better understand the evolution of this new disease and implement adequate and efficient management and prevention measures. In fact, the progression of COVID-19 disease can vary between asymptomatic disease, mild or moderate flu-like illness and pneumonia with severe respiratory failure, that often requires hospitalization in specialized wards and in severe cases in sub-intensive or intensive care unit [3]. The course of the disease varies considerably between individuals based on their demographic characteristics and previous health conditions, so it is difficult to define the prognosis for each individual. Age and sex are confirmed and well described prognostic factors, with a higher probability of developing serious illness and a higher mortality rate in older subjects and males [4, 5]. Several studies also argue that the presence of pre-existing comorbidities, such as diabetes and hypertension, is a determinant of poor prognosis [6, 7]. Furthermore, the epidemiological dynamics of COVID-19 has changed radically over the months due to, for example, variable climatic conditions [8], the presence of more or less stringent restrictions imposed by governments and the effect of vaccination [9]. From the beginning of the pandemic at the end of February 2020 until fall 2021, Italy, for example, has recorded three major waves of COVID-19 infections, interspersed with periods of less spread of the virus [10].

In light of the foregoing, it is essential to gain a better understanding of key prognostic factors of COVID-19 disease and quantify the strength of their association with the patient’s likelihood of experiencing a critical event to identify patients at high risk of clinical deterioration. Furthermore, in order to predict the evolution of the pandemic and improve its management, an analysis of the temporal trends of the disease and of the effect of the vaccination campaign on the severity of the latter is also necessary.

With this purpose, our study considers 32500 cases of SARS-CoV-2 infection diagnosed in the municipality of Bologna from February 28/2020 to October 13/2021 and through statistical tools analyzes the risk factors for hospitalization in general COVID-19 wards and in COVID-19 intensive or sub-intensive care units. Of particular interest were the age, sex and comorbidities of the patients, the vaccination status at the time of diagnosis and the calendar period of diagnosis.

## Materials and Methods

### Study design and participants

We performed a retrospective cohort study on 32500 laboratory-confirmed cases of COVID-19 in the municipality of Bologna from February 28/2020 to October 13/2021. The analysis, performed between December 2021 and January 2023, describes the progression of patients from the diagnosis of the disease to their admission to the intensive or sub-intensive care unit (both referred to as ICU in the following), passing through the eventual admission to COVID-19 general wards. An individual was considered censored after ending the disease without experiencing hospitalization.

### Data collection, setting and data organization

The data set analysed in this study was obtained with permission from the Local Health Unit (AUSL, *Azienda Unità Sanitaria Locale*) of the municipality of Bologna. It comprises various information about dates of diagnosis, hospital and ICU admission, recovery and vaccination, socio demographic characteristics (age, sex, neighborhood of residence,…) of the participants, presence of symptoms during the period of illness and comorbidities. No missing data were present. We do not have reliable and certain data on deaths. In this regard, our study stops at hospitalization in intensive care considered as the only absorbing state. In particular, in the data set provided by the AUSL of Bologna are listed all the entrances and exits from all hospital wards, COVID-19 dedicated and not, in the studied period of time, relating to the major hospitals in the municipality of Bologna, namely Sant’Orsola Hospital, Maggiore Hospital, Bellaria Hospital, Bentivoglio Hospital, Budrio Hospital, San Giovanni in Persiceto Hospital, Bazzano Hospital, Porretta Hospital, Loiano Hospital and Vergato Hospital.

For the purpose of this study, the patients were divided into eight age groups (0-19, 20-29, 30-39, 40-49, 50-59, 60-69, 70-79 and over 80) based on the ISS (*Istituto Superiore di Sanità*) guide lines [11].

Furthermore, the participants were divided into five groups according to their calendar period of diagnosis [12]:

1. Wave 1 (W1): from February 28/2020 to May 13/2020;
2. Out-wave 1 (OW1): from May 14/2020 to November 06/2020;
3. Wave 2 (W2): from November 07/2020 to February 01/2021;
4. Wave 3 (W3): from February 02/2021 to April 28/2021;
5. Out-wave 2 (OW2): from April 29/04 to October 31/2021.

These periods were identified by observing the number of daily hospitalizations in COVID-19 wards. In particular, the periods in which this number exceeds 500 are called waves, while those in which is less than 500 are named out-waves (S1 Fig).

Finally, the patients were divided into three groups according to the vaccination status at the time of diagnosis. There is a well-known delay between inoculation and artificial immunization [13], and in particular it was found that in case of SARS-CoV-2 vaccines this is about 14 days after the first dose and 7 days after the second dose for mRNA vaccines (Pfizer-BioNTech, Moderna) [14] and 14 days after the single dose of Johnson&Johnson vaccine [15]. For this reason, we considered individuals as not vaccinated (NV) up to 14 days after the first dose of mRNA vaccine or up to 14 days after the single dose of the J&J vaccine, partially vaccinated (PV) as of 14 days after their first dose of mRNA vaccine and totally vaccinated (TV) as of 7 days after their second dose of mRNA vaccine or 14 days after being vaccinated with the single dose of J&J [16, 17]. People partially vaccinated who tested positive for COVID-19 after more than 150 days from the administration of the first dose (after which immunity is indeterminate) were excluded in order to take into account potentially late second doses but, at the same time, not excessively overestimate the effectiveness of the partial immunization [16]. Furthermore, people vaccinated with Astrazeneca S.P.A. vaccine were not considered since the use of this vaccine had a troubled history in Italy in the period under analysis [18]. In particular, several people who received the first dose of Astrazeneca S.P.A vaccine were then given a second dose of an mRNA vaccine.

Therefore, we felt that it was difficult to estimate the effectiveness of the vaccine in the case of individuals who received at least one dose of Astrazeneca S.P.A vaccine and we exclude them from the analysis. Therefore the final number of patients considered is equal to 32500, i.e. the total number of COVID cases in the municipality of Bologna in the period considered (34016) minus the number of patients who received at least one dose of the Astrazeneca S.P.A vaccine (1516).

### Statistical analysis

First of all, the data have been reworked in order to make them suitable for statistical analysis. In particular, all the independent variables (sex, age group, calendar period of diagnosis, comorbidities and vaccination status) were considered as categorical variables and dummy coded, i.e. 1 if the individual presents a certain characteristics and 0 otherwise. Such pre-processing was performed using the package *pandas* v1.4.1 [19] in Python v3.8.8.

Hence, the Kaplan-Meier method was used to estimate the probability of avoiding hospitalization in ICU (considered as the event) at any time after the diagnosis of COVID-19 and in particular the difference in this probability was observed between males and females. Log rank test was used to test any difference in survival probability among the two sexes. This analysis was performed using the packages *lifelines* v0.26.4 [20] and *matplotlib* v3.5.1 for the visualization [21] in Python.

Subsequently, we built a multi-state model (Fig 1) to investigate the determinants of risk for three possible transitions that patients can experience during the disease, that is from COVID-19 diagnosis (DIAG) to hospitalization in non-ICU COVID-19 specialised wards (HOSP); from DIAG to admission to ICU (ICU) and from HOSP to ICU. A multivariate Cox’s proportional hazard model, was fit to each transition using age groups, calendar period of diagnosis, vaccination status and pathologies as predictor factors.

**Fig 1.**
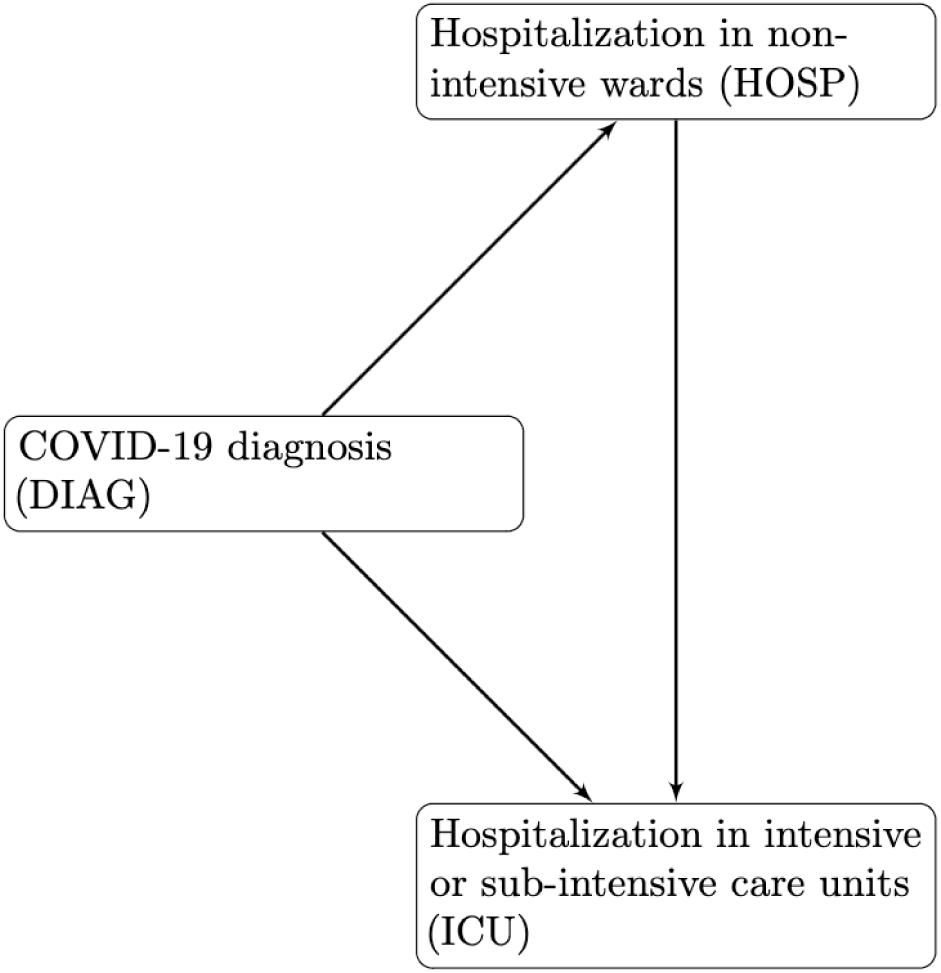
States and tran5itions for the multi-state model.

The Cox’s model is expressed by the hazard function denoted by *h(t)* (Eq (1)) which can be interpreted as the instantaneous risk of experiencing the event of interest at time *t*:

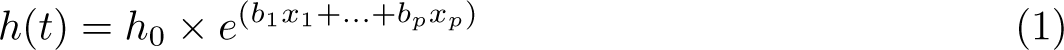

where the (*x*_1_,…,*x_p_*) is the set of predictor factors (covariates), the coefficients (*b*_1_,…,*b_p_*) measure the impact of covariates on the event realization and *h*_0_ is the baseline hazard function, that corresponds to the value of the hazard when all the *x_i_*are equal to 0. A value of *b_i_* greater than 0, or equivalently an hazard ratio HR = *e^b^^i^* greater than 1, indicates that as the value of the *i*-th covariate increases, the event hazard increases as well. On the other hand, a value of *b_i_* smaller than 0 (HR *<* 1), indicates that the *i*-th covariate is negatively associated with the event probability.

Therefore, through a multivariate Cox’s model we studied the effect of sex, age group, calendar period of diagnosis, vaccination status at the time of diagnosis and pathologies on the probability of experiencing any of the three transitions. We have chosen W2 as reference period in the analyses in order to better observe the differences between the beginning of the pandemic that caught the whole world unprepared (W1 and OW1) and a more advanced phase of the same characterized by a better knowledge of the virus and the launch of the vaccination campaign (W3 and OW2). We checked the Cox proportional hazard assumptions by Schoenfeld residuals test and plot. This statistical analysis was conducted in R v4.1.1 using the packages *survminer* v0.4.9 [22] and *survival* v3.5-5 [23]. Specifically, the multivariate Cox’s model was fit by means of the function ”coxph” of the *survival* package with the default parameters.

Given the limited number of performed hypothesis tests p-values were not adjusted for multiple testing. Furthermore, p-values less than 0.05 were considered statistically significant. In particular, we assign one star (^*^) if p *<* 0.05, two stars (^**^) if p *<* 0.01 and three stars (^***^) if p *<* 0.001.

## Results and Discussion

### Description of study cohort

Our study cohort was composed of 32500 people, who tested positive for COVID-19 between February 28/2020 and October 13/2021 in the municipality of Bologna, i.e. the total number of COVID cases in the municipality of Bologna in the period considered (34016) minus the number of patients who received at least one dose of the Astrazeneca S.P.A vaccine (1516).

Overall, 16696 (51.37%) of the participants were females, while 15804 (48.63%) were males.

Based on ISS (*Istituto Superiore di Sanité*) guide lines [11], the following age groups were considered for the analysis: 0-19, 20-29, 30-39, 40-49, 50-59, 60-69, 70-79 and over 80 years old.

Of all participants, 1497 (4.61%) have contracted COVID-19 during W1, 4090 (12.58%) during OW1, 10017 (30.82%) during W2, 12376 (38.08%) during W3 and the remaining 4520 (13.91%) during OW2.

Regarding hospital admissions, 29423 individuals (90.53%) did not require hospitalization, 2475 (7.62%) were hospitalised but did not enter ICU and 602 (1.85%) entered the ICU.

### Kaplan-Meier curves of ICU hospitalization

Table 1 shows the percentages of patients, divided by sex, age group, calendar period of diagnosis, vaccination status and comorbidities who were admitted to ICU during the infection and those of censored people, that is who left the analysis without having experienced such hospitalization. All patients were followed up from the first positive COVID-19 test until censored or ICU entry (event). Of the 602 registered events, 586 (97.34%) occurred in less than 14 days following initial COVID-19 diagnosis. The Kaplan-Meier estimation technique was used to calculate the probability of not being admitted to ICU as days pass from the diagnosis (Fig 2 - A). As expected, the overall Kaplan-Meier curve decreases rapidly during the first 14 days after COVID-19 diagnosis showing that most entries in ICU occurred during this time. Separate Kaplan-Meier curves stratified by sex (Fig 2 - B) show a statistically significant difference between males and females in the probability of not being admitted to ICU. In particular, the latter is higher in women at any time after COVID-19 diagnosis. A similar result was found in several articles [5, 24, 25], confirming male sex as a poor prognostic factor.

**Table 1.** Distribution of patients by sex, age group, calendar period of diagnosis, vaccination status and pathologies.

|  | **Infection** | **ICU** |  | **Censored** |  |
| --- | --- | --- | --- | --- | --- |
|  | **No.** | **No.** | **%** | **No.** | **%** |
| **Total** | 32500 | 602 | 1.85 | 31898 | 98.15 |
| **Sex** |  |  |  |  |  |
| Male | 15804 | 403 | 2.55 | 15401 | 97.45 |
| Female | 16696 | 199 | 1.19 | 16497 | 98.81 |
| **Age group** |  |  |  |  |  |
| 0-19 | 5521 | 5 | 0.10 | 5516 | 99.90 |
| 20-29 | 3942 | 7 | 0.18 | 3935 | 99.82 |
| 30-39 | 4688 | 27 | 0.58 | 4661 | 99.42 |
| 40-49 | 5437 | 60 | 1.10 | 5377 | 98.90 |
| 50-59 | 5461 | 115 | 2.11 | 5346 | 97.89 |
| 60-69 | 2952 | 158 | 5.35 | 2794 | 94.65 |
| 70-79 | 1833 | 146 | 7.97 | 1687 | 92.03 |
| over 80 | 2666 | 84 | 3.15 | 2582 | 96.85 |
| **Calendar period of diagnosis** |  |  |  |  |  |
| W1 | 1497 | 70 | 4.68 | 1427 | 95.32 |
| OW1 | 4090 | 85 | 2.08 | 4005 | 97.92 |
| W2 | 10017 | 179 | 1.79 | 9838 | 98.21 |
| W3 | 12376 | 236 | 1.91 | 12140 | 98.09 |
| OW2 | 4520 | 32 | 0.71 | 4488 | 99.29 |
| **Vaccination status** |  |  |  |  |  |
| NV | 30938 | 596 | 1.93 | 30342 | 98.07 |
| PV | 417 | 5 | 1.20 | 412 | 98.80 |
| TV | 1145 | 1 | 0.10 | 1144 | 99.90 |
| **Pathology** |  |  |  |  |  |
| Essential hypertension | 1380 | 71 | 5.14 | 1309 | 94.86 |
| Hypertensive heart disease | 456 | 30 | 6.58 | 426 | 93.42 |
| Diabetes mellitus | 1230 | 103 | 8.37 | 1127 | 91.63 |
| Hypercholesterolemia | 384 | 18 | 4.69 | 365 | 95.05 |
| Malignant neoplasm | 1177 | 45 | 3.82 | 1132 | 96.18 |
| Multiple sclerosis | 68 | 0 | 0.00 | 68 | 100.00 |
| Hypothyroidism | 620 | 66 | 10.65 | 554 | 89.35 |
| Glaucoma | 256 | 11 | 4.30 | 245 | 95.70 |
| Toxic diffuse goiter | 62 | 1 | 1.61 | 61 | 98.39 |
| Epilepsy | 78 | 3 | 3.85 | 75 | 96.15 |
| Heart and pulmonary circulation diseases | 872 | 59 | 6.77 | 813 | 93.23 |
| Chronic kidney failure | 184 | 14 | 7.61 | 170 | 92.39 |
| Celiac disease | 170 | 0 | 0.00 | 170 | 100.00 |
| Asthma | 357 | 8 | 2.24 | 349 | 97.76 |
| Rheumatoid arthritis | 108 | 9 | 8.33 | 99 | 91.67 |
| Chronic Hepatitis | 301 | 15 | 4.98 | 286 | 95.02 |
| Embolism and thrombosis of other veins | 145 | 18 | 12.41 | 127 | 87.59 |
| Psoriasis | 52 | 0 | 0.00 | 52 | 100.00 |
| Hashimoto's thyroiditis | 320 | 3 | 0.94 | 317 | 99.06 |
| Ulcerative colitis | 106 | 1 | 0.94 | 105 | 99.06 |
| Parkinson's disease | 30 | 2 | 6.66 | 28 | 93.34 |
| Pituitary dwarfism | 58 | 0 | 0.00 | 58 | 100.00 |
| Endometriosis (III - IV ASRM stage) | 40 | 0 | 0.00 | 40 | 100.00 |

**Fig 2.**
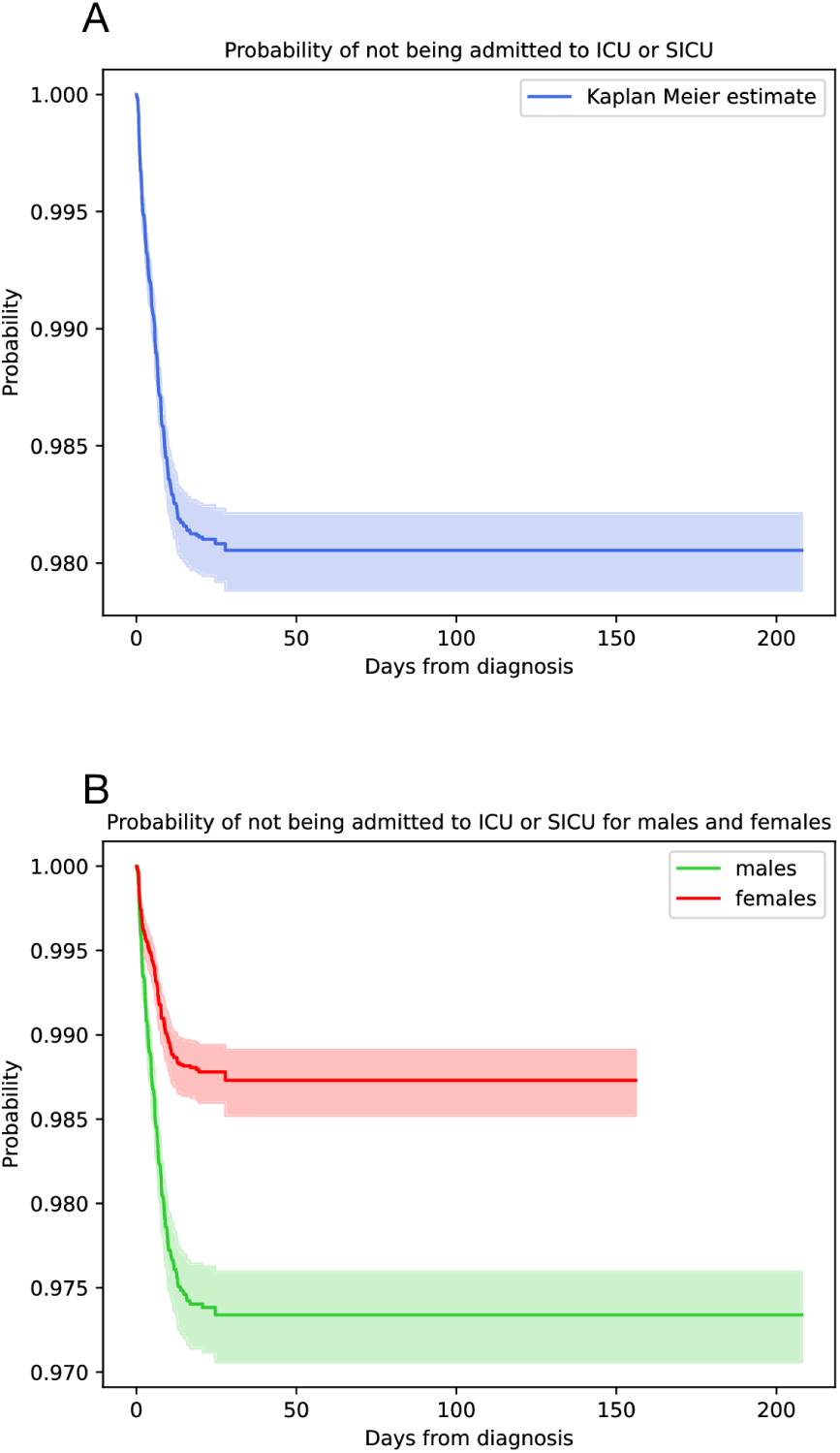
Kaplan-Meier curves. Overall (A) and stratified by sex (B) estimate of the probability of not being admitted to ICU during COVID-19 infection.

### Multi-state Cox’s Regression model

A multivariate and multi-state Cox’s proportional hazard model was fitted to identify the factors associated to the risk of experiencing three potential transitions during the course of the disease. The scheme of the model’s events and transitions is represented in Fig 1. Three transitions were studied: from COVID-19 diagnosis to hospitalization in any COVID-19 ward except ICU (DIAG*→*HOSP); from COVID-19 diagnosis to hospitalization in ICU (DIAG*→*ICU); from admission to any non-ICU COVID-19 ward to admission to ICU (HOSP*→*ICU). The percentages of patients divided by gender, age, period of diagnosis, vaccination status and pathologies who have experienced each of these transitions are shown in Table 2.

**Table 2.** Distribution of patients by sex, age group, calendar period of diagnosis, vaccination status, pathologies and transition.

|  | DIAG | DIAG→HOSP |  | DIAG→ICU |  | HOSP→ICU |  |
| --- | --- | --- | --- | --- | --- | --- | --- |
|  | No. | No. | % of DIAG | No. | % of DIAG | No. | % of HOSP |
| **Total** | 32500 | 2831 | 8.71 | 246 | 0.76 | 356 | 12.58 |
| **Sex** |  |  |  |  |  |  |  |
| Male | 15804 | 1572 | 9.95 | 152 | 0.96 | 251 | 15.97 |
| Female | 16696 | 1259 | 7.54 | 94 | 0.56 | 105 | 8.34 |
| **Age group** |  |  |  |  |  |  |  |
| 0-19 | 5521 | 11 | 0.20 | 4 | 0.07 | 1 | 9.09 |
| 20-29 | 3942 | 48 | 1.22 | 4 | 0.10 | 3 | 6.25 |
| 30-39 | 4688 | 133 | 2.84 | 12 | 0.26 | 15 | 11.28 |
| 40-49 | 5437 | 352 | 6.47 | 31 | 0.57 | 29 | 8.24 |
| 50-59 | 5461 | 512 | 9.38 | 47 | 0.86 | 68 | 13.28 |
| 60-69 | 2952 | 528 | 17.89 | 63 | 2.13 | 95 | 17.99 |
| 70-79 | 1833 | 517 | 28.21 | 51 | 2.78 | 95 | 18.38 |
| over 80 | 2666 | 730 | 27.38 | 34 | 1.28 | 50 | 6.85 |
| **Calendar period of diagnosis** |  |  |  |  |  |  |  |
| W1 | 1497 | 436 | 29.12 | 28 | 1.87 | 42 | 9.63 |
| OW1 | 4090 | 323 | 7.90 | 44 | 1.08 | 41 | 12.69 |
| W2 | 10017 | 700 | 6.99 | 72 | 0.72 | 107 | 15.29 |
| W3 | 12376 | 1137 | 9.19 | 89 | 0.72 | 147 | 12.93 |
| OW2 | 4520 | 235 | 5.20 | 13 | 0.29 | 19 | 8.09 |
| **Vaccination status** |  |  |  |  |  |  |  |
| NV | 30938 | 2746 | 8.88 | 242 | 0.78 | 354 | 12.89 |
| PV | 417 | 38 | 9.11 | 3 | 0.72 | 2 | 5.26 |
| TV | 1145 | 47 | 4.10 | 1 | 0.10 | 0 | 0.00 |
| **Pathology** |  |  |  |  |  |  |  |
| Essential hypertension | 1380 | 331 | 23.99 | 21 | 1.52 | 50 | 15.11 |
| Hypertensive heart disease | 456 | 97 | 21.27 | 13 | 2.85 | 17 | 17.53 |
| Diabetes mellitus | 1230 | 326 | 26.50 | 41 | 3.33 | 62 | 19.02 |
| Hypercholesterolemia | 384 | 84 | 21.88 | 4 | 1.04 | 15 | 17.86 |
| Malignant neoplasm | 1177 | 219 | 18.61 | 16 | 1.36 | 29 | 13.24 |
| Multiple sclerosis | 68 | 11 | 16.18 | 0 | 0.00 | 0 | 0.00 |
| Hypothyroidism | 620 | 66 | 10.65 | 3 | 0.48 | 12 | 18.18 |
| Glaucoma | 256 | 57 | 22.27 | 3 | 1.17 | 8 | 14.04 |
| Toxic diffuse goiter | 62 | 3 | 4.84 | 1 | 1.61 | 0 | 0.00 |
| Epilepsy | 78 | 6 | 7.69 | 1 | 1.28 | 2 | 33.33 |
| Heart and pulmonary circulation diseases | 872 | 200 | 22.94 | 24 | 2.75 | 35 | 17.50 |
| Chronic kidney failure | 184 | 74 | 40.22 | 5 | 2.72 | 9 | 12.16 |
| Celiac disease | 170 | 5 | 2.94 | 0 | 0.00 | 0 | 0.00 |
| Asthma | 357 | 41 | 11.48 | 4 | 1.12 | 4 | 9.76 |
| Rheumatoid arthritis | 108 | 22 | 20.37 | 4 | 3.70 | 5 | 22.73 |
| Chronic Hepatitis | 301 | 49 | 16.28 | 9 | 2.99 | 6 | 12.24 |
| Embolism and thrombosis of other veins | 145 | 42 | 28.97 | 10 | 6.90 | 8 | 19.05 |
| Psoriasis | 52 | 8 | 15.38 | 0 | 0.00 | 0 | 0.00 |
| Hashimoto's thyroiditis | 320 | 19 | 5.94 | 1 | 0.31 | 2 | 10.53 |
| Ulcerative colitis | 106 | 14 | 13.21 | 0 | 0.00 | 1 | 7.14 |
| Parkinson's disease | 30 | 10 | 33.33 | 0 | 0.00 | 2 | 20.00 |
| Pituitary dwarfism | 58 | 1 | 1.72 | 0 | 0.00 | 0 | 0.00 |
| Endometriosis (III - IV ASRM stage) | 40 | 2 | 5.00 | 0 | 0.00 | 0 | 0.00 |

Specifically, the survival model was used to analyse the effects of age, sex, calendar period of diagnosis, vaccination status at the time of diagnosis and pathologies on the risk of experiencing any of those transition. The results of this model can be useful both immediately and in the future for making public health decisions. For example, to identify patients most at risk of hospitalization who need a priority for vaccination and to improve the use of resources in hospitals, such as oxygen tanks. Furthermore, these results could be used to prevent overcrowding in intensive care units, which has been one of the major problems in managing the pandemic in Italy [26], by identifying the factors that increase the risk of such recovery especially among patients already hospitalized in order to be able to treat them adequately before their conditions worsen.

Overall, our results show that the Cox’s model fits the data well. Table 3 shows in fact that, considering a 5-folds cross-validation, a concordance index of 0.82 is obtained on the test set for the DIAG*→*HOSP transition, a concordance index of 0.80 for the DIAG*→*ICU transition, and a concordance index of 0.64 for the HOSP*→*ICU transition.

**Table 3.** Mean concordance indices and standard errors assigned on the training set and test set in a 5-fold cross validation.

| Transition | Training set | Test set |
| --- | --- | --- |
| DIAG→HOSP | 0.820 (0.001) | 0.817 (0.004) |
| DIAG→ICU | 0.827 (0.003) | 0.798 (0.011) |
| HOSP→ICU | 0.687 (0.002) | 0.642 (0.011) |

For what concerns the partial effects of the covariates in the model, a common result of all three transitions analyzed is an increased risk of experiencing them associated with males compared to females (HR 1.65 95% CI 1.53 - 1.79 for DIAG*→*HOSP transition; HR 1.81 95% CI 1.39 - 2.37 for DIAG*→*ICU transition; HR 1.92 95% CI 1.50 - 2.45 for HOSP*→*ICU transition). This finding is in line with many studies in which a significantly higher risk of mortality, hospitalisation and ICU admission was found to be linked with male sex [24, 25].

### Transition DIAG*→*HOSP

Table 4 shows the Cox’s model results for the DIAG→HOSP transition.

**Table 4.**
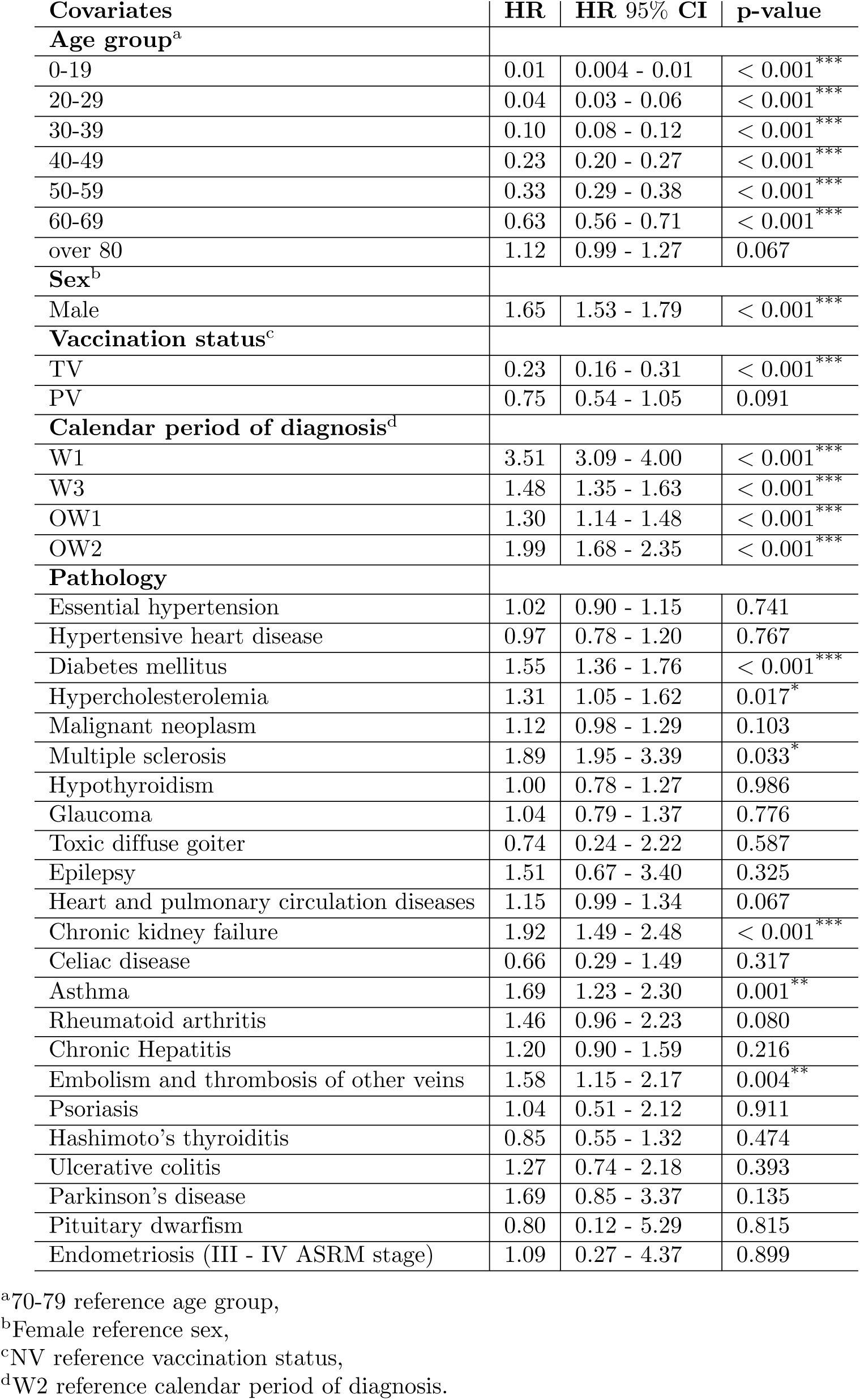
Cox’s model results for DIAG→HOSP transition.

The hazard ratio generally increases with age. However, no significant increase is observed in the risk of being admitted to any COVID-19 ward for those over 80 compared to the reference age group 70-79 (HR 1.12 95% CI 0.99 - 1.27). A similar result was obtained in a Spanish study examining the association between age and the risk of COVID-19 diagnosis, hospitalization and death which shows that the risk of hospitalization after a diagnosis of COVID-19 peaked, for both sexes, around the age of 75 and then decreased slightly for older ages [27].

Taking the not vaccinated status (NV) as baseline, it results that being partially immunized does not imply a significant change in the risk of experiencing DIAG*→*HOSP transition (HR 0.75 95% CI 0.54 - 1.05). On the other hand, the totally vaccinated status (TV) is associated with a significant reduction in the hazard of hospitalization (HR 0.23 95% CI 0.16 - 0.31).

As regards the effect of the calendar period of diagnosis, having contracted the disease during W1 or W3 implied a significant greater risk of being hospitalized in a non-ICU COVID-19 ward than during W2 (3.51 95% CI 3.09 - 4.00 for W1; 1.15 95% CI 1.35 - 1.63 for W3). Despite some differences in the definition of the three waves, our results are comparable with those obtained from an analysis of the COVID-19 outcomes of 4 million inhabitants of Northwest Italy during the first three waves of the pandemic [28]. In particular, this study confirmed a greater risk of hospitalization in both non-ICU and ICU wards of an infected person in the first wave compared to the second wave. This may have been due to the improvements in COVID-19 treatment following the first wave of the epidemic, such as the use of more effective pharmacological strategies, and the widening of access to test for COVID-19 that was initially restricted to the most severely ill patients. However, as regards what obtained for W3, it is difficult to find an explanation to this result since no significant changes in the management and treatment of the pandemic are present between W2 and W3 periods. Probably, this finding is due to factors beyond our knowledge such as, for example, the availability of beds in hospitals dedicated to infected people during the two waves which can affect the number of hospitalized patients.

Among all the pathologies taken into consideration, diabetes (HR 1.55 95% CI 1.36 - 1.76), hypercholesterolemia (HR 1.31 95% CI 1.05 - 1.62), multiple sclerosis (HR 1.89 95% CI 1.95 - 3.39), chronic kidney failure (HR 1.92 95% CI 1.49 - 2.48), asthma (HR 1.69 95% CI 1.23 - 2.30) and embolism (HR 1.58 95% CI 1.15 - 2.17) were significantly associated with a increased risk of hospitalization. These results are in line with several studies that have observed a significantly higher risk of non-ICU hospitalization among patients with diabetes, chronic kidney failure and asthma [24, 29, 30].

### Transition DIAG→ICU

Table 5 reports the hazard ratios resulted from the Cox’s model for the DIAG*→*ICU transition.

**Table 5.** Cox’s model results for DIAG→ICU transition.

| Covariates | HR | HR 95% CI | p-value |
| --- | --- | --- | --- |
| **Age group<sup>a</sup>** |  |  |  |
| 0-19 | 0.03 | 0.01 - 0.09 | < 0.001*** |
| 20-29 | 0.04 | 0.02 - 0.01 | < 0.001*** |
| 30-39 | 0.11 | 0.06 - 0.21 | < 0.001*** |
| 40-49 | 0.24 | 0.15 - 0.39 | < 0.001*** |
| 50-59 | 0.34 | 0.22 - 0.53 | < 0.001*** |
| 60-69 | 0.83 | 0.56 - 1.22 | 0.342 |
| over 80 | 0.57 | 0.36 - 0.89 | 0.013* |
| **Sex<sup>b</sup>** |  |  |  |
| Male | 1.81 | 1.39 - 2.37 | < 0.001*** |
| **Vaccination status<sup>c</sup>** |  |  |  |
| TV | 0.10 | 0.01 - 0.73 | 0.023* |
| PV | 1.15 | 0.36 - 3.68 | 0.820 |
| **Calendar period of diagnosis<sup>d</sup>** |  |  |  |
| W1 | 2.30 | 1.47 - 3.60 | < 0.001*** |
| W3 | 1.06 | 0.77 - 1.44 | 0.729 |
| OW1 | 1.64 | 1.13 - 2.39 | 0.010* |
| OW2 | 0.99 | 0.53 - 1.87 | 0.981 |
| **Pathology** |  |  |  |
| Essential hypertension | 0.79 | 0.49 - 1.28 | 0.338 |
| Hypertensive heart disease | 1.50 | 0.84 - 2.68 | 0.174 |
| Diabetes mellitus | 2.43 | 1.65 - 3.59 | < 0.001*** |
| Hypercholesterolemia | 0.56 | 0.21 - 1.47 | 0.237 |
| Malignant neoplasm | 0.91 | 0.54 - 1.53 | 0.707 |
| Multiple sclerosis | - | - | - |
| Hypothyroidism | 0.46 | 0.15 - 1.48 | 0.194 |
| Glaucoma | 0.65 | 0.20 - 2.09 | 0.470 |
| Toxic diffuse goiter | 3.05 | 0.51 - 18.36 | 0.224 |
| Epilepsy | 2.39 | 0.33 - 17.49 | 0.390 |
| Heart and pulmonary circulation diseases | 1.65 | 1.06 - 2.56 | 0.028* |
| Chronic kidney failure | 1.44 | 0.55 - 3.73 | 0.457 |
| Celiac disease | - | - | - |
| Asthma | 1.75 | 0.64 - 4.77 | 0.276 |
| Rheumatoid arthritis | 3.90 | 1.43 - 10.63 | 0.008** |
| Chronic Hepatitis | 2.39 | 1.23 - 4.65 | 0.010* |
| Embolism and thrombosis of other veins | 4.94 | 2.57 - 9.51 | < 0.001*** |
| Psoriasis | - | - | - |
| Hashimoto's thyroiditis | 0.55 | 0.08 - 3.97 | 0.553 |
| Ulcerative colitis | - | - | - |
| Parkinson's disease | - | - | - |
| Pituitary dwarfism | - | - | - |
| Endometriosis (III - IV ASRM stage) | - | - | - |

<sup>a</sup>70-79 reference age group,<sup>b</sup>Female reference sex,<sup>c</sup>NV reference vaccination status,<sup>d</sup>W2 reference calendar period of diagnosis.

The hazard increases with age but only up to 60-69. In fact, the 60-69 group does not show a significantly different hazard compared to 70-79 (HR 0.83 95% CI 0.56 - 1.22). For what concerns the over 80 group, instead, a slightly significant lower hazard is observed with respect to the reference (HR 0.57 95% CI 0.36 - 0.89). These findings are in line with what obtained in a Swedish study, that is a decrease of the risk in ICU hospitalization for ages older than 60-69 [29]. This could be due to the fact that several elderly people may have died even before being admitted to intensive care or to the fact that in some critical periods of the pandemic, priority was given to younger people who needed admission to intensive care [31]. Furthermore, the fact that the elderly were the most protected and least exposed category of people during the pandemic as the Italian government recommended them to limit contacts as much as possible [32], could have influenced the decrease in the risk of admission to intensive care for these people.

Concerning the effect of the vaccination status, no significant difference in the hazard of being directly hospitalized in ICU after diagnosis was observed among partially (PV) and non vaccinated (NV) patients (HR 1.15 95% CI 0.36 - 3.68). As for those who have contracted the disease after completing the vaccination cycle (TV), instead, a slightly significant decrease in risk was found (HR 0.10 95% CI 0.01 - 0.73).

As regards the influence of the calendar period of diagnosis, W1 is associated with an increased hazard to experience the DIAG*→*ICU transition compared to W2 (HR 2.30 95% CI 1.47 - 3.60). On the other hand, no significant difference in the risk of hospitalization in ICU between patients infected by SARS-CoV-2 during W3 and those tested positive for COVID-19 during W2 was observed (HR 1.06 95% CI 0.77 - 1.44). Although the definitions of the different waves do not match perfectly as they are based on arbitrary criteria for each study, our results are fairly in line with other studies in which changes in ICU admission and mortality have been analyzed over time [28, 33]. In particular, in each of these studies, as in ours, a reduction in the risk of serious illness was observed in the second wave compared to the first one. As already discussed, this result is the effect of the development and improvement of the clinical management of the new disease after the first few months. Furthermore, the absence of difference in the risk of ICU admission found between W2 and W3 may be justified, as said before, by the fact that there were no significant changes in the management of the pandemic and treatment of the disease between the two periods.

Being affected by diabetes (HR 2.43 95% CI 1.65 - 3.59), heart or pulmonary circulation disease (HR 1.65 95% CI 1.06 - 2.56), rheumatoid arthritis (HR 3.90 95% CI 1.43 - 10.63), chronic hepatitis (HR 2.39 95% CI 1.23 - 4.65) or embolism (HR 4.94 95% CI 2.57 - 9.51), was found to be associated with a significantly higher risk of being admitted to ICU. The same result regarding diabetes was obtained in a nationwide Swedish study [29].

### Transition HOSP→ICU

Table 6 shows the results of the Cox’s model for the HOSP*→*ICU transition.

**Table 6.** Cox’s model results for HOSP***→***ICU transition.

| Covariates | HR | HR 95% CI | p-value |
| --- | --- | --- | --- |
| **Age group<sup>a</sup>** |  |  |  |
| 0-19 | 0.50 | 0.06 - 4.01 | 0.514 |
| 20-29 | 0.38 | 0.12 - 1.23 | 0.106 |
| 30-39 | 0.64 | 0.36 - 1.17 | 0.147 |
| 40-49 | 0.45 | 0.29 - 0.70 | < 0.001*** |
| 50-59 | 0.77 | 0.55 - 1.08 | 0.126 |
| 60-69 | 1.09 | 0.81 - 1.48 | 0.562 |
| over 80 | 0.40 | 0.28 - 0.58 | < 0.001*** |
| **Sex<sup>b</sup>** |  |  |  |
| Male | 1.92 | 1.50 - 2.45 | < 0.001*** |
| **Vaccination status<sup>c</sup>** |  |  |  |
| TV | - | - | - |
| PV | 0.48 | 0.12 - 1.99 | 0.315 |
| **Calendar period of diagnosis<sup>d</sup>** |  |  |  |
| W1 | 0.53 | 0.37 - 0.76 | < 0.001*** |
| W3 | 0.94 | 0.72 - 1.21 | 0.613 |
| OW1 | 0.77 | 0.53 - 1.12 | 0.179 |
| OW2 | 0.67 | 0.39 - 1.13 | 0.133 |
| **Pathology** |  |  |  |
| Essential hypertension | 1.12 | 0.81 - 1.56 | 0.495 |
| Hypertensive heart disease | 1.21 | 0.72 - 2.95 | 0.478 |
| Diabetes mellitus | 1.18 | 0.87 - 1.59 | 0.286 |
| Hypercholesterolemia | 1.44 | 0.85 - 2.43 | 0.178 |
| Malignant neoplasm | 0.82 | 0.54 - 1.25 | 0.361 |
| Multiple sclerosis | - | - | - |
| Hypothyroidism | 1.62 | 0.84 - 3.15 | 0.152 |
| Glaucoma | 1.01 | 0.50 - 2.07 | 0.973 |
| Toxic diffuse goiter | - | - | - |
| Epilepsy | 1.82 | 0.44 - 7.51 | 0.408 |
| Heart and pulmonary circulation diseases | 1.13 | 0.79 - 1.62 | 0.496 |
| Chronic kidney failure | 0.77 | 0.38 - 1.53 | 0.447 |
| Celiac disease | - | - | - |
| Asthma | 0.79 | 0.28 - 2.23 | 0.663 |
| Rheumatoid arthritis | 2.17 | 0.79 - 5.93 | 0.132 |
| Chronic Hepatitis | 0.89 | 0.40 - 2.00 | 0.780 |
| Embolism and thrombosis of other veins | 1.70 | 0.83 - 3.50 | 0.147 |
| Psoriasis | - | - | - |
| Hashimoto's thyroiditis | 1.30 | 0.31 - 5.46 | 0.724 |
| Ulcerative colitis | 0.45 | 0.07 - 3.10 | 0.420 |
| Parkinson's disease | 0.91 | 0.22 - 3.74 | 0.898 |
| Pituitary dwarfism | - | - | - |
| Endometriosis (III - IV ASRM stage) | - | - | - |

<sup>a</sup>70-79 reference age group,<sup>b</sup>Female reference sex,<sup>c</sup>NV reference vaccination status,<sup>d</sup>W2 reference calendar period of diagnosis.

Only few patients in the age ranges 0-19, 20-29, or 30-39 were present in the initial state (i.e. few patients of these age groups were hospitalized in non-ICU wards; see Table 2 for details), making the hazard ratios related to those groups not robust. For this reason, we focused our discussion only on older ages. A significantly lower risk was observed for patients aged 40-49 and over 80 compared to 70-79 (HR 0.45 95% CI 0.29 - 0.70 for 40-49; HR 0.40 95% CI 0.28 - 0.58 for over 80), while no significant difference in transition risk was noticed for ages 50-59 and 60-69 (HR 0.77 95% CI 0.55 - 1.08 for 50-59; HR 1.09 95% CI 0.81 - 1.48 for 60-69). Likewise, in a study from Kuwait [34], it was found that the probability of ICU admission among hospitalized patients was highest at around age 70 and then decreases for older ages. As already discussed, these findings could be the effect, at least in part, of the rationing of healthcare resources during the critical phases of the pandemic with younger patients being prioritized to receive hospital care and intensive services if needed [31].

For what concerns the influence of the vaccination status, as for the transitions previously analyzed, being partially vaccinated (PV) does not imply any change in the risk of experiencing HOSP*→*ICU transition (HR 0.48 95% CI 0.12 - 1.99). On the other hand, no person among those totally vaccinated (TV) at the time of diagnosis was transferred to ICU from another COVID-19 ward. Therefore, consistent with an official ISS report dated September 2021 that studies the impact of vaccination with mRNA vaccines on the risk of COVID-19 infection, hospitalization and death by analyzing data from the National Vaccination Registry and the COVID-19 Integrated Surveillance System [35], we observed that the completion of the vaccination cycle significantly reduces the risk of COVID-19-related hospitalisation. Our results are also in line with a Norwegian study in which it was observed that fully vaccinated hospitalized patients had a significantly lower risk of ICU admission than unvaccinated ones [36].

As regards the effect of the calendar period of diagnosis, a significantly lower risk of HOSP*→*ICU transition was observed during W1 than during W2 (HR 0.53 95% CI 0.37 - 0.76), while during the other calendar periods of diagnosis there is no significant difference in the same hazard compared to the reference.

None of the pathologies considered was associated with a significant increase in the risk of moving to ICU from a non-ICU ward. This result is in contradiction with what emerged in some studies in which for example patients with diabetes [37] and asthma [24] are significantly more at risk of experiencing this transition.

### Study limitations

The results of our study should be interpreted taking into account some limitations. The major constrain of this study is the lack of available data on deaths among SARS-CoV-2 infected people. Therefore, it was not possible for us to analyze the factors that influence mortality but only those associated with the risk of developing a severe form of the disease that requires hospitalization in intensive care. This makes the multi-state model incomplete as we have been forced to consider ICU as an absorbing state when in reality it is possible to get out of it by dying or passing to a state other than death, such as discharge, recovery or admission to another non-intensive ward.

Furthermore, this study only concerns the municipality of Bologna, hence the results obtained cannot be generalized to other municipalities or to all of Italy or other countries. We believe that an accurate comparison with other studies should also take into account specific factors of each territory such as, for example, the socio-economic conditions of the study cohort, the criteria of virus testing in the study setting, the availability of, the availability of beds in local hospitals for COVID-19 patients and in general the management of the pandemic by the competent authorities during the study period.

Another limitation of this study is that the criterion used to define the waves is arbitrary and based only on daily hospitalizations in the municipality of Bologna during the study period. However, our definition is quite in line with other Italian studies [28, 33] and national reports [10].

In addition, we did not take into account the circulation of several variants of SARS-CoV-2 over the study period, whose different virulence may have affected the rate of hospitalization and the mortality.

Finally, although studies have found that COVID-19 vaccines only start to provide significant protection after one week after the second dose of mRNA vaccines or two weeks after the single J&J vaccine dose [14, 15], the immune response triggered by vaccination is gradual and variable between individuals. Therefore, our categorization of patients by vaccination status at the time of COVID-19 diagnosis, although supported by several articles [16, 17], may be too simplistic not taking into account this gradual growth and variability of immunity.

## Conclusion

As the novel coronavirus disease has become a major public health event due to its rapid transmission and large-scale spread, it is necessary to prepare in advance about the risks of serious illness to prevent pandemic degeneration based on local demographics, the current situation of medical resources and the progress of the vaccination campaign. For this reason, our study aimed to investigate the factors associated with the risk of developing a severe form of the disease requiring hospitalization in both intensive and non-intensive wards. Using a multi-state model, the study revealed that older age, male sex, having contracted the disease during the first wave, and being unvaccinated or partially vaccinated are poor prognostic factors for COVID-19. The multi-state model approach made it possible to study the transition of patients between diagnosis, admission to non-intensive wards, and admission to intensive care unit in a more detail way than using the two-state model. In addition, we identified some diseases associated with an increased risk of ICU and non-ICU hospitalization, including diabetes, asthma and chronic kidney failure. In conclusion, our findings suggest that to limit the number of severe cases of COVID-19 and consequently the overcrowding of hospitals, the completion of the vaccination cycle and a more careful monitoring and prioritization in the care of the elderly and of people with certain specific comorbidities are essential.

## Supporting information

S1 Fig. Graphical representation of the number of monthly hospitalizations in any COVID-19 ward during the study period in the hospitals of the municipality of Bologna. (TIFF)

S2 Fig. Distribution of patients by transitions for each vaccination status. (TIFF)

## Ethics statement

Ethical approval for the study was obtained from the University of Bologna’s Ethical Committee (approval number 283066, 5 October 2021). The data set was previously anonymized, and the authors did not have access to any information that could have been used to identify the individual patients involved in the study. In compliance with Italian Data protection Authority [38], no informed consent was required for carrying out observational studies for scientific research purposes.

## Data Availability Statement

Data of the present study were provided from the Local Health Unit (AUSL, *Azienda Unità Sanitaria Locale*) of the municipality of Bologna, Italy, and the authors do not have the right to share them. In order to gain access to the data, contact Local Health Unit of the municipality of Bologna, Italy.

## Data Availability

Data of the present study were provided from the Local Health Unit (AUSL, Azienda 385 Unit`a Sanitaria Locale) of the municipality of Bologna, Italy, and the authors do not 386 have the right to share them. In order to gain access to the data, contact Local Health 387 Unit of the municipality of Bologna, Italy.

## Acknowledgments

We acknowledge the Bologna MODELS4COVID Study Group of the University of Bologna and the National Institute for Nuclear Physics (INFN): Armando Bazzani, Valerio Carelli, Paolo Tubertini, Luca Clissa, Stefano Diciotti, Enrico Lunedei, Michela Milano, Luca Palmerini, Daniel Remondini, Giulia Roli, Michele Scagliarini, Roberto Spighi, Vincenzo Vagnoni, Antonio Zoccoli, Lorenzo Chiari.

## References

1. Worobey M, Levy JI, Malpica Serrano L, Crits-Christoph A, Pekar JE, Goldstein SA et al. The Huanan Seafood Wholesale Market in Wuhan was the early epicenter of the COVID-19 pandemic. Science. 2022 Aug 26;377(6609):951–959. doi: 10.1126/science.abp8715.

2. COVID-19 situation update for the EU/EEA [cited 2022 October 08]. In: European Centre for Disease Prevention and Control [Internet]. Available from: https://www.ecdc.europa.eu/en/cases-2019-ncov-eueea.

3. Clinical Spectrum of SARS-CoV-2 Infection [cited 2022 October 08]. In: National Institutes of Health [Internet]. Available from: https://www.covid19treatmentguidelines.nih.gov/overview/clinical-spectrum/.

4. Parohan M, Yaghoubi S, Seraji A, Hassan Javanbakht M, Sarraf P, Djalali M. Risk factors for mortality in patients with Coronavirus disease 2019 (COVID-19) infection: a systematic review and meta-analysis of observational studies. The Aging Male. 2020 Jun 08;23(5):1416–1424. doi: 10.1080/13685538.2020.1774748.

5. Pijls BG, Jolani S, Atherley A, Derckx RT, Dijkstra JIR, Franssen GHL et al. Demographic risk factors for COVID-19 infection, severity, ICU admission and death: a meta-analysis of 59 studies. BMJ Open. 2021;11:e044640. doi: 10.1136/bmjopen-2020-044640.

6. Guo W, Li M, Dong Y, Zhou H, Zhang Z, Tian C et al. Diabetes is a risk factor for the progression and prognosis of COVID-19. Diabetes Metab Res Rev. 2020 Mar 31;36(7):e3319. doi: 10.1002/dmrr.3319.

7. Pranata R, Lim MA, Huang I, Raharjo SB, Lukito AA. Hypertension is associated with increased mortality and severity of disease in COVID-19 pneumonia: A systematic review, meta-analysis and meta-regression. J Renin Angiotensin Aldosterone Syst. 2020 Apr-Jun;21(2):1470320320926899. doi: 10.1177/1470320320926899.

8. Liu X, Huang J, Li C, Zhao Y, Wang D, Huang Z et al. The role of seasonality in the spread of COVID-19 pandemic. Environ Res. 2021 Apr;195: 110874. doi: 10.1016/j.envres.2021.110874.

9. Liu Q, Qin C, Liu M, Liu J. Effectiveness and safety of SARS-CoV-2 vaccine in real-world studies: a systematic review and meta-analysis. Infect Dis Poverty. 2021;10: 132. doi: 10.1186/s40249-021-00915-3.

10. Coronavirus in Italia: dati, infografiche e mappe [cited 17 October 2022]. In: Sky tg24 [Internet]. Available from: https://tg24.sky.it/cronaca/approfondimenti/coronavirus-italia-contagi.

11. Coronavirus [cited 21 September 2022]. In: EpiCentro-Istituto Superiore di Sanità [Internet]. Available from: https://www.epicentro.iss.it/coronavirus/.

12. Zeleke AJ, Moscato S, Miglio R, Chiari L. Length of Stay Analysis of COVID-19 Hospitalizations Using a Count Regression Model and Quantile Regression: A Study in Bologna, Italy. Int J Environ Res Public Health. 2022 Feb 16;19(4):2224. doi: 10.3390/ijerph19042224.

13. Pollard AJ, Bijker EM. A guide to vaccinology: from basic principles to new developments. Nat Rev Immunol. 2021 Feb; 21(2):83–100. doi: 10.1038/s41577-020-00479-7.

14. Polack FP, Thomas SJ, Kitchin N, Absalon J, Gurtman A, Lockhart S et al. C4591001 Clinical Trial Group. Safety and Efficacy of the BNT162b2 mRNA Covid-19 Vaccine. N Engl J Med. 2020 Dec 31;383(27):2603–2615. doi: 10.1056/NEJMoa2034577.

15. FDA Briefing Document. Janssen Ad26.COV2.S Vaccine for the Prevention of COVID-19. 2021 February 26 [cited 2022 October 20]. Available from: https://www.fda.gov/media/146217/download.

16. Cocchio S, Zabeo F, Facchin G, Piva N, Furlan P, Nicoletti M et al. The Effectiveness of a Diverse COVID-19 Vaccine Portfolio and Its Impact on the Persistence of Positivity and Length of Hospital Stays: The Veneto Region’s Experience. Vaccines (Basel). 2022 Jan 11; 10(1):107. doi: 10.3390/vaccines10010107.

17. Tartof SY, Slezak JM, Fischer H, Hong V, Ackerson BK, Ranasinghe ON, et al. Effectiveness of mRNA BNT162b2 COVID-19 vaccine up to 6 months in a large integrated health system in the USA: a retrospective cohort study. Lancet. 2021 Oct 16; 398(10309):1407–1416. doi: 10.1016/S0140-6736(21)02183-8.

18. Pignataro M. Grey areas and uncertainties: the AstraZeneca case in Italy. Lex-Atlas: Covid-19. 2021 Aug 10 [cited 08 October 2022]. Available from: https://lexatlas-c19.org/grey-areas-and-uncertainties-the-astrazeneca-case-in-italy/.

19. McKinney W. Data Structures for Statistical Computing in Python. Proceedings pf 9th Python in Science Conference. 2010; 56–61. doi: 10.25080/Majora-92bf1922-00a.

20. Davidson-Pilon C. lifelines: survival analysis in Python. Journal of Open Source Software. 2019;4(40):1317. doi: 10.21105/joss.01317.

21. Hunter JD. Matplotlib: A 2D graphics environment. Computing in Science & Engineering. 2007;9(3):90–95. doi: 10.1109/MCSE.2007.55.

22. Kassambara A, Kosinski M, Biecek P. survminer: Drawing Survival Curves using ’ggplot2’. R package version 0.4.9. 2021 [cited 03 April 2023]. Available from: https://CRAN.R-project.org/package=survminer.

23. Therneau T. A Package for Survival Analysis in R. R package version 3.5-5. 2023 [cited 03 April 2023]. Available from: https://CRAN.R-project.org/package=survival.

24. Bennett KE, Mullooly M, O’Loughlin M, Fitzgerald M, O’Donnell J, O’Connor L et al. Underlying conditions and risk of hospitalisation, ICU admission and mortality among those with COVID-19 in Ireland: A national surveillance study. Lancet Reg Health Eur. 2021 Jun;5:100097. doi: 10.1016/j.lanepe.2021.100097.

25. Sisó-Almirall A, Kostov B, Mas-Heredia M, Vilanova-Rotllan S, Sequeira-Aymar E, Sans-Corrales M, et al. Prognostic factors in Spanish COVID-19 patients: A case series from Barcelona. PLoS One. 2020 Aug 21;15(8):e0237960. doi: 10.1371/journal.pone.0237960.

26. Dettaglio terapie intensive in Italia [cited 17 October 2022]. In: Statistiche coronavirus [Internet]. Available from: https://statistichecoronavirus.it/coronavirus-italia/terapie-intensive/.

27. Burn E, Tebé C, Fernandez-Bertolin S, Aragon M, Recalde M, Roel E, et al. The natural history of symptomatic COVID-19 during the first wave in Catalonia. Nat Commun. 2021 Feb 3;12(1):777. doi: 10.1038/s41467-021-21100-y.

28. D’Arminio Monforte A, Tavelli A, Bai F, Tomasoni D, Falcinella C, Caramello V et al. Improvements throughout the Three Waves of COVID-19 Pandemic: Results from 4 Million Inhabitants of North-West Italy. J Clin Med. 2022 Jul 25;11(15):4304. doi: 10.3390/jcm11154304.

29. Bergman J, Ballin M, Nordström A, Nordström P. Risk factors for COVID-19 diagnosis, hospitalization, and subsequent all-cause mortality in Sweden: a nationwide study. Eur J Epidemiol. 2021;36:287–298. doi:10.1007/s10654-021-00732-w.

30. Ko JY, Danielson ML, Town M, Derado G, Greenlund KJ, Kirley PD, et al. COVID-NET Surveillance Team. Risk Factors for Coronavirus Disease 2019 (COVID-19)-Associated Hospitalization: COVID-19-Associated Hospitalization Surveillance Network and Behavioral Risk Factor Surveillance System. Clin Infect Dis. 2021 Jun 1;72(11):e695–e703. doi: 10.1093/cid/ciaa1419.

31. Privitera G. Italian doctors on coronavirus frontline face tough calls on whom to save. POLITICO. 2020 Mar 09 [cited 2022 October 21]. Available from: https://www.politico.eu/article/coronavirus-italy-doctors-tough-calls-survival/.

32. Covid-19, le raccomandazioni per le persone anziane [cited 2022 October 21]. In: Ministero della Salute [Internet]. Available from: https://www.salute.gov.it/portale/nuovocoronavirus/dettaglioNotizieNuovoCoronavirus.jsp?id=4172&lingua=italiano&menu=notizie&p=dalministero.

33. Giacomelli A, Ridolfo AL, Pezzati L, Oreni L, Carrozzo G, Beltrami M et al. Mortality rates among COVID-19 patients hospitalised during the first three waves of the epidemic in Milan, Italy: A prospective observational study. PLoS One. 2022 Apr 11;17(4):e0263548. doi: 10.1371/journal.pone.0263548.

34. Kipourou DK, Leyrat C, Alsheridah N, Almazeedi S, Al-Youha S, Jamal MH et al. Probabilities of ICU admission and hospital discharge according to patient characteristics in the designated COVID-19 hospital of Kuwait. BMC Public Health. 2021;21:799. doi: 10.1186/s12889-021-10759-z.

35. Istituto Superiore di Sanità. Impact of COVID-19 vaccination on the risk of SARS-CoV-2 infection and hospitalization and death in Italy. Report n. 4 of 2021 Sept 30 [cited 2022 October 17]. Available from: https://www.iss.it/documents/20126/0/report_on_vaccine_effectiveness_Italy+%281%29.pdf/53d71dc2-c8c5-24c1-3467-705a8587a339?t=1633529045681.

36. Whittaker R, Bråthen Kristofferson A, Valcarcel Salamanca B, Seppälä E, Golestani K, Kvåle R, et al. Length of hospital stay and risk of intensive care admission and in-hospital death among COVID-19 patients in Norway: a register-based cohort study comparing patients fully vaccinated with an mRNA vaccine to unvaccinated patients. Clin Microbiol Infect. 2022 Jun;28(6):871–878. doi: 10.1016/j.cmi.2022.01.033.

37. Kim L, Garg S, O’Halloran A, Whitaker M, Pham H, Anderson EJ et al. Risk Factors for Intensive Care Unit Admission and In-hospital Mortality Among Hospitalized Adults Identified through the US Coronavirus Disease 2019 (COVID-19)-Associated Hospitalization Surveillance Network (COVID-NET). Clin Infect Dis. 2021 May 4;72(9):e206–e214. doi: 10.1093/cid/ciaa1012.

38. Garante per la Protezione dei Dati Personali. General Authorisation to Process Personal Data for Scientific Research Purposes - 1 March 2012 [1884019] [cited 2023 April 20]. Available from: https://www.garanteprivacy.it/home/docweb/-/docweb-display/docweb/1884019.

