## Supplementary figures and images for "Risk Factors for Admission into COVID-19 General Wards, Sub-Intensive and Intensive Care Units among SARS-CoV-2 Positive Subjects in the Municipality of Bologna, Italy"

### S1 Fig

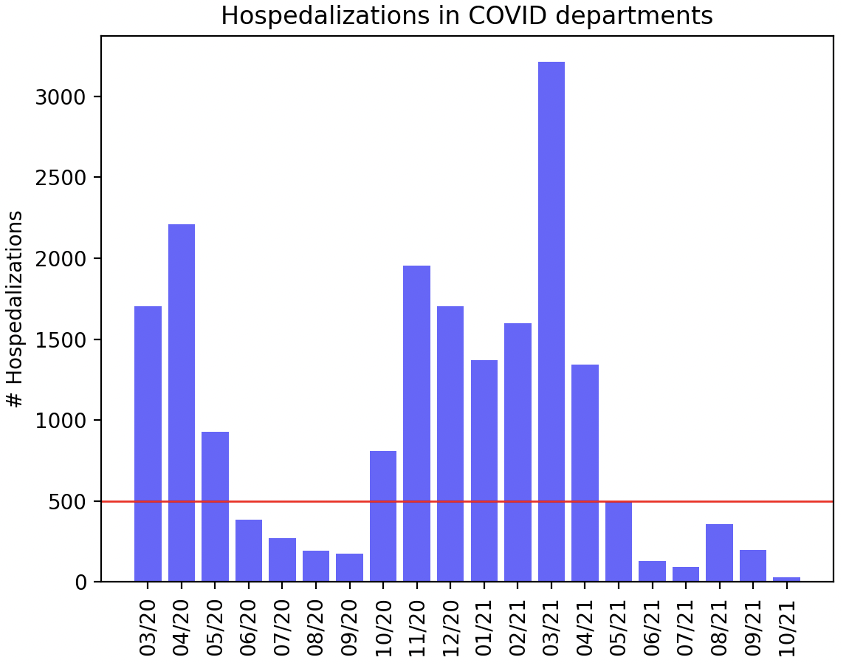

### S2 Fig

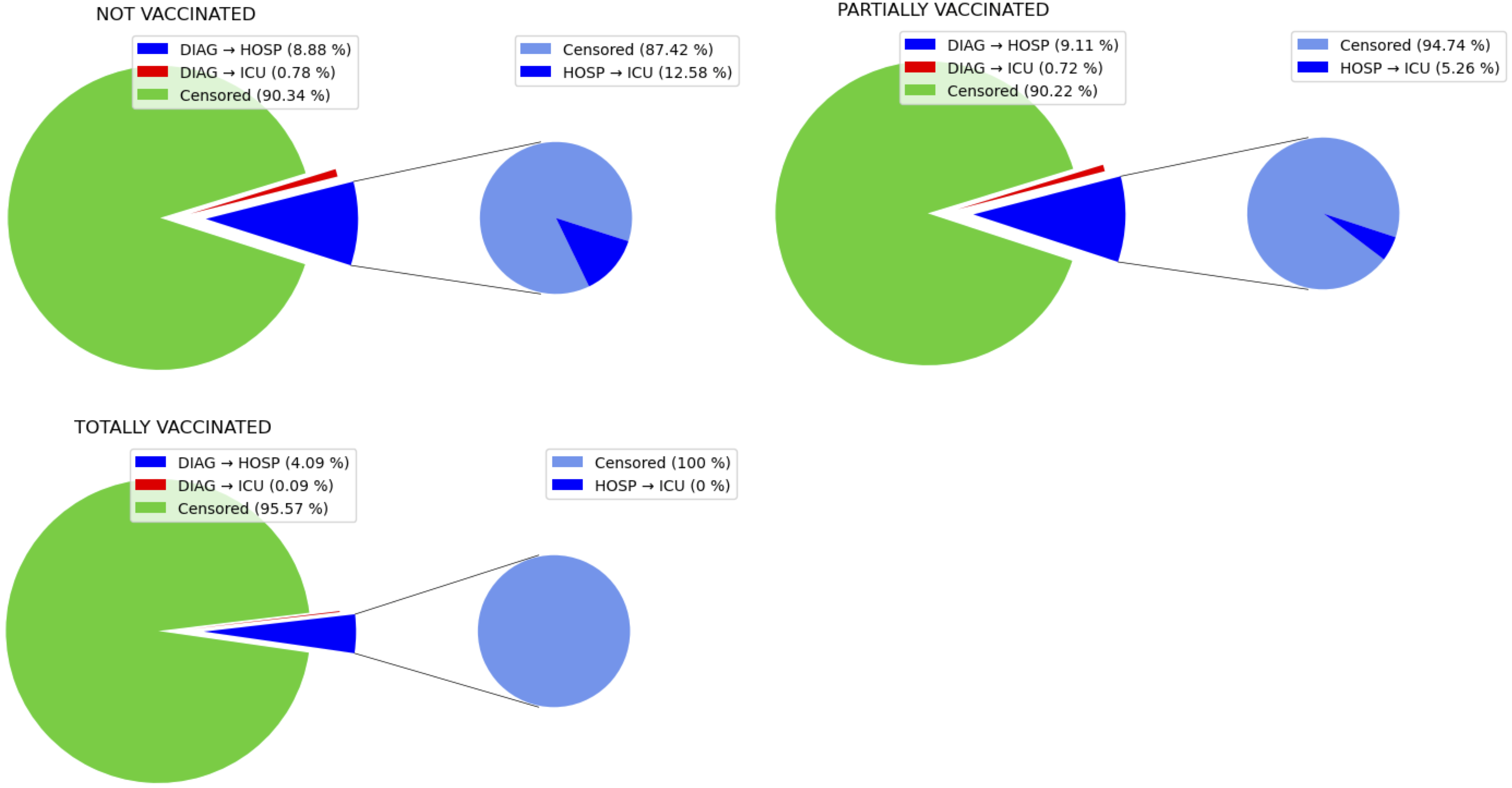
